# Comparison of two T cell assays to evaluate T cell responses to SARS-CoV-2 following vaccination in naïve and convalescent healthcare workers

**DOI:** 10.1101/2022.02.05.22270447

**Authors:** Eloise Phillips, Sandra Adele, Tom Malone, Alexandra Deeks, Lizzie Stafford, Susan L. Dobson, Ali Amini, Donal Skelly, David Eyre, Katie Jeffery, Christopher P. Conlon, Christina Dold, Ashley Otter, Silvia D’Arcangelo, Lance Turtle, PITCH Consortium, Paul Klenerman, Eleanor Barnes, Susanna J. Dunachie

## Abstract

**Background:** T cell responses to SARS-CoV-2 following infection and vaccination are less characterised than antibody responses, due to a more complex experimental pathway.

**Methods:** We measured T cell responses in 108 healthcare workers (HCWs) in an observational cohort study, using the commercialised Oxford Immunotec T-SPOT Discovery SARS-CoV-2 assay (OI T-SPOT) and the PITCH ELISpot protocol established for academic research settings.

**Results:** Both assays detected T cell responses to SARS-CoV-2 spike, membrane and nucleocapsid proteins. Responses were significantly lower when reported by OI T-SPOT than by PITCH ELISpot. Four weeks after two doses of either Pfizer/BioNTech BNT162b or ChAdOx1 nCoV-19 AZD1222 vaccine, the responder rate was 63% for OI T-SPOT Panels1+2 (peptides representing SARS-CoV-2 spike protein excluding regions present in seasonal coronaviruses), 69% for OI T-SPOT Panel 14 (peptides representing the entire SARS-CoV-2 spike), and 94% for the PITCH ELISpot assay. The two OI T-SPOT panels correlated strongly with each other showing that either readout quantifies spike-specific T cell responses, although the correlation between the OI T-SPOT panels and the PITCH ELISpot was moderate.

**Conclusion:** The standardisation, relative scalability and longer interval between blood acquisition and processing are advantages of the commercial OI T-SPOT assay. However, the OI T-SPOT assay measures T cell responses at a significantly lower magnitude compared to the PITCH ELISpot assay, detecting T cell responses in a lower proportion of vaccinees. This has implications for the reporting of low-level T cell responses that may be observed in patient populations and for the assessment of T cell durability after vaccination.

## Introduction

With rapid roll-out of SARS-CoV-2 vaccinations across global healthcare systems, measurement of immune responses in both partially and fully vaccinated individuals is desirable for comparison of vaccination regimens, evaluation of immunocompromised individuals, monitoring responses to emerging variants of concern and for determining the need for boosters. Surveillance of immune responses can guide COVID-19 vaccine rollout schemes to reduce risk of infection and disease severity, and enable better allocation of healthcare resources. A number of cross-sectional and prospective cohort studies of healthcare workers (HCWs) have been established in order to monitor immune responses in individuals with vaccine- and/or infection-acquired immunity [1-3].

Monitoring of antibody responses to SARS-CoV-2, especially neutralising antibodies, receives the most focus from researchers and policy makers. Serum for antibody assays is relatively easy to collect and store, binding antibodies can be measured at scale by automated platforms, and correlations with protection at a population level have been observed for binding [4] and neutralising antibodies [5-8]. Nevertheless, T cells are a key arm of the immune response, orchestrating the antigen-specific adaptive response to infection including optimal production of antibodies from B cells, as well as having cytotoxic properties against virally infected cells. There is some evidence in macaque models and humans that the T cell response is important in defence against SARS-CoV-2 [9, 10]. T cell responses are maintained after boosting with a second vaccine dose [11], and the anti-spike (anti-S) T cell response following vaccination with Pfizer/BNT162b2 does not correlate precisely with anti-S IgG antibody response [11]. Importantly, unlike the humoral response, the T cell response to SARS-CoV-2 is minimally impacted by mutations in the alpha, beta, gamma and delta variants of concern [11, 12], and 75-85% preserved against the omicron variant [13-19]. Therefore quantifying the T cell response to SARS-CoV-2 is important but such monitoring is largely restricted to dedicated research centres due the technical expertise required to isolate cells from fresh blood within hours of blood draw, and the relative complexity of assays.

The *ex vivo* interferon-gamma enzyme-linked absorbent spot (IFN-γ ELISpot) assay is a common workhorse assay used to measure antigen-specific T cell responses. Specifically, ELISpot measures secreted cytokines at the single-cell level from peripheral blood, and by stimulating these cells with specific antigens of interest, T cell responses to these antigens can be monitored. The main advantage afforded by ELISpot is its sensitivity, which exceeds cytokine flow cytometry assays [20] and is up to 200 times greater than ELISA [21]. The assay takes two days to perform, using skilled technicians. For this reason, collaboration with third-party diagnostic companies is becoming increasingly common in clinical research, especially for SARS-CoV-2. Such collaborations enable efficient data output and facilitate rapid research, the results of which can accordingly be used to inform clinical practice and patient management.

The PITCH Study (Protective Immunity from T-cells in Healthcare workers) is a UK multi-centre prospective, observational cohort study in Oxford, Birmingham, Liverpool, Newcastle and Sheffield which investigates T-cell responses in both vaccine- and/or infection-acquired immunity to SARS-CoV-2 infection [22]. We used this setting to evaluate the use of the proprietary Oxford Immunotec T-SPOT Discovery SARS-CoV-2 assay [23] (OI T-SPOT), alongside T cell measurement by our in-house IFNγ ELISpot assay using the PITCH protocol, that has been harmonised across the five PITCH centres.

This study sought to compare use of the Oxford Immunotec T-SPOT Discovery SARS-CoV-2 assay (OI T-SPOT) in reporting T cell responses specific to SARS-CoV-2 spike and structural proteins, with our in-house PITCH ELISpot. Both assays are based on the ELISpot technique. The OI T-SPOT assay, which is an example of a commercial interferon-gamma release assay (IGRA), was introduced by Oxford Immunotec 15 years ago as a diagnostic test for *Mycobacterium tuberculosis* (T-SPOT.*TB* test). A kit form with pre-coated plates and anti-IFN-γ antibodies is available for immunology laboratories. This initial technology was optimised to enumerate *M. tuberculosis*-sensitised T cells by measuring IFN-γ secreted from CD4 and CD8 T cells in response to antigens from *M. tuberculosis* (Reviewed [24]). As the SARS-CoV-2 pandemic spread, this technology was subsequently adapted to allow SARS-CoV-2-specific T cells to be enumerated. The PITCH ELISpot protocol has origins at the Jenner Institute [25] where it was initially developed to enumerate T cell responses to malaria by measuring IFN-γ secretion to a pre-erythrocytic malaria antigen, thrombospondin-related adhesion protein (TRAP). This technique was later altered with SARS-CoV-2 antigens [26] before being harmonised for PITCH across 5 UK laboratories [22]. Here we compared these two SARS-CoV-2 ELISpot assays to demonstrate the utility of the OI T-SPOT assay.

## Materials and Methods

### Study design and participants

In this prospective, observational, cohort study, we sampled participants at one PITCH centre in Oxford, UK. HCWs were enrolled in the OPTIC study (GI Biobank Study 16/YH/0247, approved by the research ethics committee (REC) at Yorkshire & The Humber - Sheffield Research Ethics Committee on 29 July 2016, and amended for the OPTIC study on 8 June 2020. Healthy men and women aged 18 years and older were recruited, with all working HCWs eligible and outreach activities by the team to encourage participation from a wide range of age, sex, ethnicity and work role.

Previous SARS-CoV-2 infection status was defined in HCWs based on documented PCR and/or baseline anti-S and anti-N serology results from the Abbott platform at Oxford University Hospital NHS Foundation Trust prior to vaccination. All participants received either the BNT162b Pfizer/BioNTech vaccine or the ChAdOx1 nCoV-19 AZD1222 vaccine. We aimed to recruit a minimum of 20 infection-naïve and 20 previously infected per group by 28 days after the second vaccine dose, to demonstrate the use of both assays.

### Oxford Immunotec T-SPOT Discovery SARS-CoV-2 Assay

6ml of sodium heparinised whole blood per participant was shipped to Oxford Immuntec, Abingdon, UK under a commercial contract. Oxford Immunotec is independent of University of Oxford. Samples were typically shipped the same day with a few samples sent after overnight storage but within 24 hours of blood draw. According to OI information, peripheral blood mononuclear cells (PBMCs) were isolated from the whole blood and 250,000 PBMCs were added per well in the T-SPOT Discovery SARS-CoV-2 kit, an ELISPOT assay modified to measure SARS-CoV-2-specific T cell responses [27]. Each well contains an optimised antigen pool, including SARS-CoV-2 structural proteins, which stimulates T cells *in vitro* allowing their response to individual SARS-CoV-2 proteins to be measured. Peptide regions of high homology to other endemic coronaviruses were removed. Alongside negative and positive controls, a total 5 SARS-CoV-2 pools are used; S1 diagnostic (Panel 1), S2 diagnostic (Panel 2), M, NP and total spike (Panel 14), ensuring the maximal breadth of the T cell response is investigated. Sequence details of the peptides were undisclosed. Results received from OI were multiplied by 4 to report here as spot-forming units (SFU) per 10^6^ peripheral blood mononuclear cells (PBMCs).

### In-house PITCH ELISpot

The PITCH frozen ELISpot Standard Operating Procedure has been published previously [22]. PBMCs cryopreserved in fetal bovine serum and 10% DMSO were thawed in a water bath at 37°C and added dropwise to 9ml R0 (RPMI media supplemented with L-Glutamine and 10mM penicillin/streptomycin) at room temperature (RT) and then centrifuged at 400g for 5 minutes. Cell pellets were then resuspended and washed in 5ml of Rab10 (filtered R0 supplemented with 10% human serum) at RT and centrifuged again. These pellets were then resuspended in 5ml Rab10 supplemented with DNase and allowed to rest for 2-3h in an incubator at 37°C, 5% CO_2_, 95% humidity. Interferon-gamma (IFN-γ) ELISpot assays were performed using the Human IFN-γ ELISpot Basic kit (Mabtech 3420-2A). MultiScreen-I 96 ELISpot plates (Millipore, MAIPS4510) were coated overnight at 4°C or for 3-8h at RT with the capture antibody (clone 1-D1K) at 5ug/ml in PBS. Coated plates were subsequently washed twice with R0 and then blocked with 100uL/well of Rab10 for 1/2-8h at RT or 8-48h at 4°C. Rested cells were centrifuged and resuspended in 1ml Rab10 for counting on Muse™ Cell Analyser or Bio-Rad TC10™ Automated Cell Counter. After blocking, overlapping peptide pools (18-mers with 10 amino acid overlap, Mimotopes) representing spike (S1, S2), membrane (M) and nucleocapsid (NP) SARS-CoV-2 proteins were added to 200,000 PBMCs/well at a final concentration of 2ug/ml for 16-18h. S1 and S2 were added in separate test wells, M and NP were combined in a singular test well. Assays were performed in triplicate. EBV, influenza and tetanus toxoid peptide pools (2ug/ml, Proimmune PX-CEFT peptide pool) and concanavalin A (ConA) were used as positive controls, along with negative control wells (DMSO in Rab10). After overnight peptide stimulation, plates were washed 7 times with 100-200uL/well PBS-0.05% Tween and then incubated for 2-4h at RT with 50uL/well of 1ug/ml biotinylated detection antibody (clone 7-B6-1) diluted in PBS. Plates were then washed again as above and incubated for 1-2h with 50uL/well of 1ug/ml streptavidin-ALP diluted in PBS. After a final wash, spots were detected by adding 50uL/well of filtered RT BCIP/NBT stock and incubating for 5 minutes in the dark. Colour development was stopped by removal of BCIP and rinsing with cold water. Plates were air-dried for at least two nights and subsequently read on the CTL immunocapture *Cellular Technology Limited, Shaker Heights, Ohio, USA) using the Smartcount® settings. The mean spots of the negative control wells were subtracted from the test wells, and then multiplied by 5 to give antigen specific responses expressed as spot-forming units (SFU)/10^6^ PBMCs. Total spike responses were defined by adding S1 and S2 responses together. The PITCH protocol as described uses 3.2 million PBMC.

### Serological assays

Anti-spike (S) and anti-nucleocapsid (N) antibodies were measured using the Roche Elecsys® Anti-SARS-CoV-2 S and Roche Elecsys® Anti-SARS-CoV-2 N assays at the Public Health England (now United Kingdom Health Security Agency) Laboratories at Porton Down, UK. The Roche S assay is reported in units per millilitre (U/ml), which are standardised 1:1 to the WHO Binding antibody units/ml (BAU/ml). Seroconversion is defined for S as a response equal to or greater than 0.8 U/ml, and for N as a response equal or greater than 1.0 COI.

### Statistical analysis

Data was analysed in GraphPad Prism 9.1.2. Non-parametric tests were used to assess significance between data sets as non-Gaussian distribution was assumed. For matched samples, Wilcoxon’s test was used to compare two groups and Friedman’s test was used to compare three or more groups, accounting for multiple comparisons. For unmatched samples, Mann Whitney test was used to compare two groups and Kruskal Wallis test was used to compare three or more groups. For analysing correlations, two-tailed non-parametric Spearman correlation was performed. Two-tailed p values were reported with less than 0.05 considered significant.

## Results

### Human Participants

108 participants, including both SARS-CoV-2 infection-naïve (n=83) and previously infected (n=25) HCWs in Oxford, UK, were included in the study where matched data from OI T-SPOT and the PITCH ELISpot assays were available. Participants were sampled just before their second dose of vaccine which was a median 9.86, interquartile range (IQR) 6.6-11 weeks after their first dose (1 dose + 10 weeks), and again a median 4.3, IQR 4-4.6 weeks after the second dose (2 dose + 4 weeks). Pre-vaccination samples at baseline were available for a limited number of participants, but without matched results for OI T-SPOT and the PITCH ELISpot assays. All sampling was between December 2020 and July 2021. Demographic details of the participants are shown in **Table 1**. Anti-S and anti-N binding antibodies measured by Roche are shown in **Supplementary Figure 1**, with all participants seroconverting to anti-spike positivity four weeks after the second dose of vaccine (range 380-50,395 AU/ml).

**Table 1.** Characteristics of healthcare workers included in the study

|  | All | Naïve | Previously Infected |
| --- | --- | --- | --- |
| **Dosing Interval** |  |  |  |
| Days, Median (IQR) | 69 (46-77) | 69 (36.5-77) | 66 (63-77) |
| Days, Range | 17-92 | 17-92 | 19-90 |
| Weeks, Median | 9.86 | 9.86 | 9.43 |
| **Days Post V2** |  |  |  |
| Median (IQR) | 30 (28-32) | 30 (28-32.8) | 29.5 (26-31.3) |
| **N** | **108** | **83** | **25** |
| Female, N (%) | 68 (63%) | 48 (58%) | 20 (80%) |
| Male, N (%) | 40 (37%) | 35 (42%) | 5 (20%) |
| Mean Age | 35.14 | 34.74 | 36.44 |
| Age in years, Median (IQR) | 33 (24-42.5) | 33 (23-42.8) | 35 (28-42) |
| Age Range | 21-66 | 21-66 | 21-63 |
| **Infection Status, N (%)** |  |  |  |
| Naïve | 83 (77%) | 83 (100%) | 0 (0%) |
| Previous SARS-CoV-2 | 25 (23%) | 0 (0%) | 25 (100%) |
| **Ethnicity (Self Reported\*), N(% of known)** |  |  |  |
| White* | 90 (86%) | 68 (85%) | 22 (88%) |
| Asian* | 10 (10%) | 8 (10%) | 2 (8%) |
| Black* | 0 (0%) | 0 (0%) | 0 (0%) |
| Other* | 5 (5%) | 4 (5%) | 1 (4%) |
| Unreported | 3 (3%) | 3 (4%) | 0 (0%) |

### T cell responses to SARS-CoV-2 spike and structural proteins measured by commercialised OI T-SPOT and in-house PITCH ELISpot

T cell responses to spike antigens after vaccination were detected by both OI T-SPOT and PITCH ELISpot assays (**Figure 1A, Table S1**), with higher responses recorded by the PITCH assay. At 2 dose + 4 weeks, median spike-specific T cell responses in the naïve cohort measured by OI Panel 1+2 and OI Panel 14 were 28 (IQR 16-64) and 40 (IQR 16-96) SFU/10^6^ PBMCs, respectively, 6.0- and 4.2-fold lower than PITCH total spike (median=167, IQR 75-284 SFU/10^6^ PBMCs). Median responses were numerically lower for the OI Panel 1 and 2 (where peptides representing regions in the spike protein of high homology to other endemic coronaviruses had been removed) compared to OI Panel 14 (peptides representing the full spike protein) although there was no statistically significant difference between the two.

**Figure 1.**
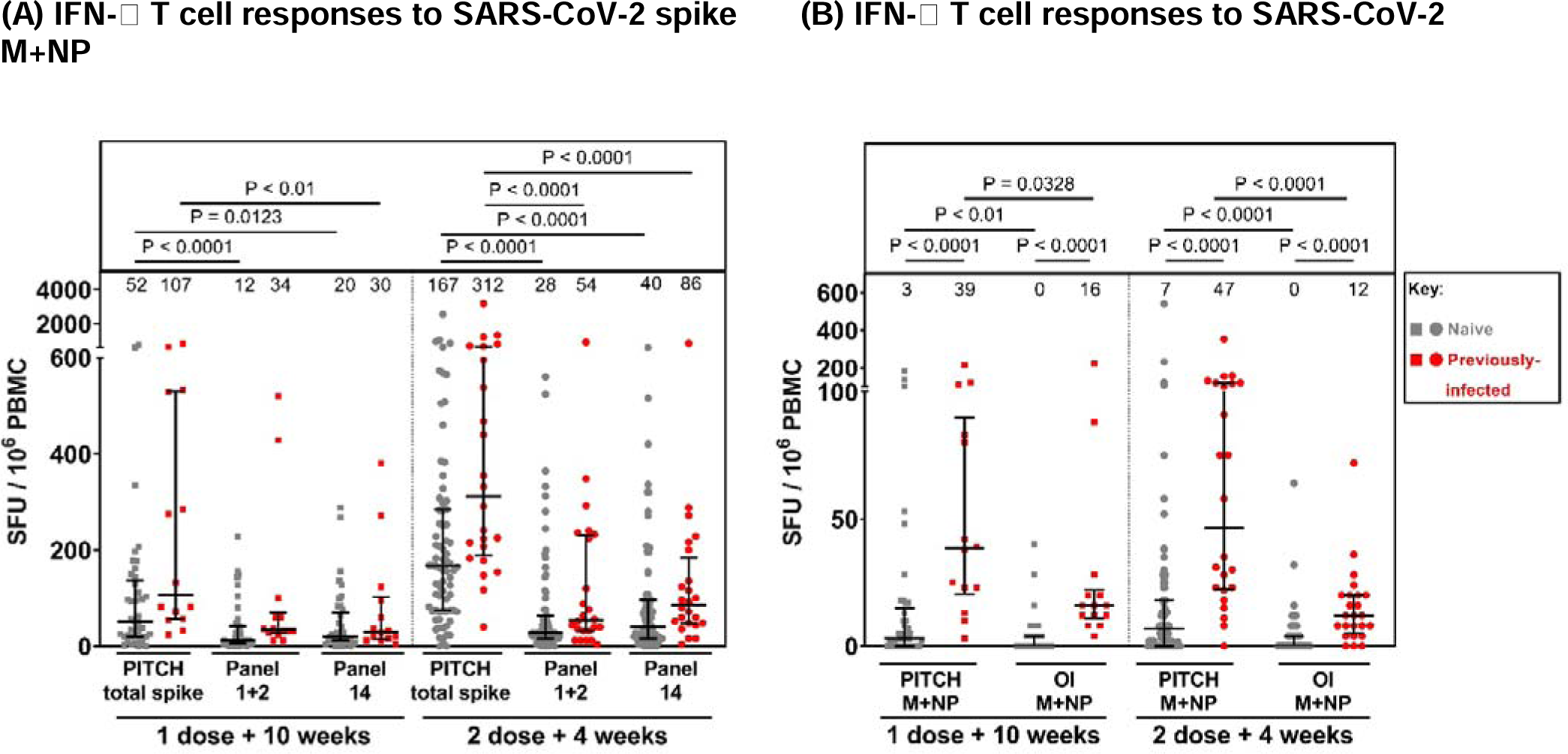
Comparison of T cell responses to SARS-CoV-2 spike and structural proteins measured by in-house PITCH ELISpot and Oxford Immunotec T-SPOT assay. (A) T cell responses to SARS-CoV-2 spike in naïve and previously-infected healthcare workers reported by three panels: PITCH total spike (S1+S2), Oxford Immunotec Panel 1+2 (diagnostic S1+S2) and Oxford Immunotec Panel 14 (total spike). Friedman test was used for statistical analysis between the 3 panels. (B) T cell responses to SARS-CoV-2 membrane (M) and nucleocapsid (NP) in naïve and previously-infected healthcare workers reported by PITCH ELISpot and Oxford Immunotec (OI) T-SPOT assay. Wilcoxon test was used for statistical analysis between samples matched across both assays, and Mann-Whitney test was used to compare naïve and previously-infected T cell responses within subgroups. (A-B) T cell responses are quantified by spot-forming units per 10^6^ peripheral blood mononuclear cells (PBMCs). Healthcare workers received phlebotomy 10 weeks post 1^st^ dose (1 dose + 10 weeks) and/or 4 weeks post 2^nd^ dose (2 dose + 4 weeks). All samples are matched across both assays (3 panels for spike and 2 for M+NP T cell responses). At 1 dose + 10 weeks, n=41 for naïve samples and n=14 for previously-infected samples. At 2 dose + 4 weeks, n=75 for naïve samples and n=24 for previously infected samples. Infection status at time of first vaccine, as defined by available PCR and serology data: grey symbols = naïve HCWs and red symbols = HCWs previously infected with SARS-CoV-2. Median T cell responses are stated immediately above each column and marked by a horizontal line on each column, and interquartile range is represented by error bars.

In SARS-CoV-2 infection-naïve HCWs at dose 2 + 4 weeks, the median T cell response to SARS-CoV-2 M+NP is 0 (IQR 0-4) SFU/10^6^ PBMCs in OI T-SPOT and 7 (IQR 0-18) SFU/10^6^ PBMCs in PITCH ELISpot (**Figure 1B**). As expected, both assays report significantly higher responses in previously-infected cohorts compared to naïve samples at each timepoint. Only previously-infected participants are expected to have T cell responses specific to SARS-CoV-2 structural proteins as natural infection involves exposure to the whole SARS-CoV-2 proteome including M+NP. Vaccination with either BNT162b Pfizer/BioNTech or ChAdOx1 nCoV-19 AZD1222 involves exposure to only SARS-CoV-2 spike protein, so naïve participants should be negative for T cell responses to SARS-CoV-2 non-spike proteins. However, some participants characterised as ‘naïve’ may have been exposed to SARS-CoV-2 without symptoms or seroconversion. Overall, our findings support the use of both assays for identifying differences in these responses between cohorts based on infection status.

### Correlation between SARS-CoV-2 specific T cell responses measured by commercialised OI T-SPOT and in-house PITCH ELISpot

To further evaluate the use of OI T-SPOT, correlations between spike-specific T cell responses reported by OI Panel 1+2, Panel 14, and PITCH total spike, were determined at 1 dose + 10 weeks (**Figure 2A-C**) and 2 dose + 4 weeks (**Figure 2E-G**). The observed correlation between OI spike panels (Panel 1+2 and Panel 14) and PITCH total spike is low to moderate at dose 1 + 10 weeks (**Figure 2A-B**) and dose 2 + 4 weeks (**Figure 2E-F**), with the strongest correlation being between OI Panel 14 and PITCH total spike at dose 2 + 4 weeks (r=0.55) and the lowest between OI Panel 1+2 and PITCH total spike at dose 2 + 4 weeks (r=0.47). As expected, Panel 1+2 and Panel 14 on OI correlate strongly at both timepoints (r=0.84 and =0.85, respectively, **Figures 2C & 2G**). Moreover, as correlations with PITCH total spike are comparable between OI Panel 1+2 and OI Panel 14, this suggests that either readout can be used to quantify T cell responses to SARS-CoV-2 spike. Weak correlations were observed for T cell responses to M+NP reported by OI T-SPOT and PITCH ELISpot (**Figure 2D & 2H**).

**Figure 2.**
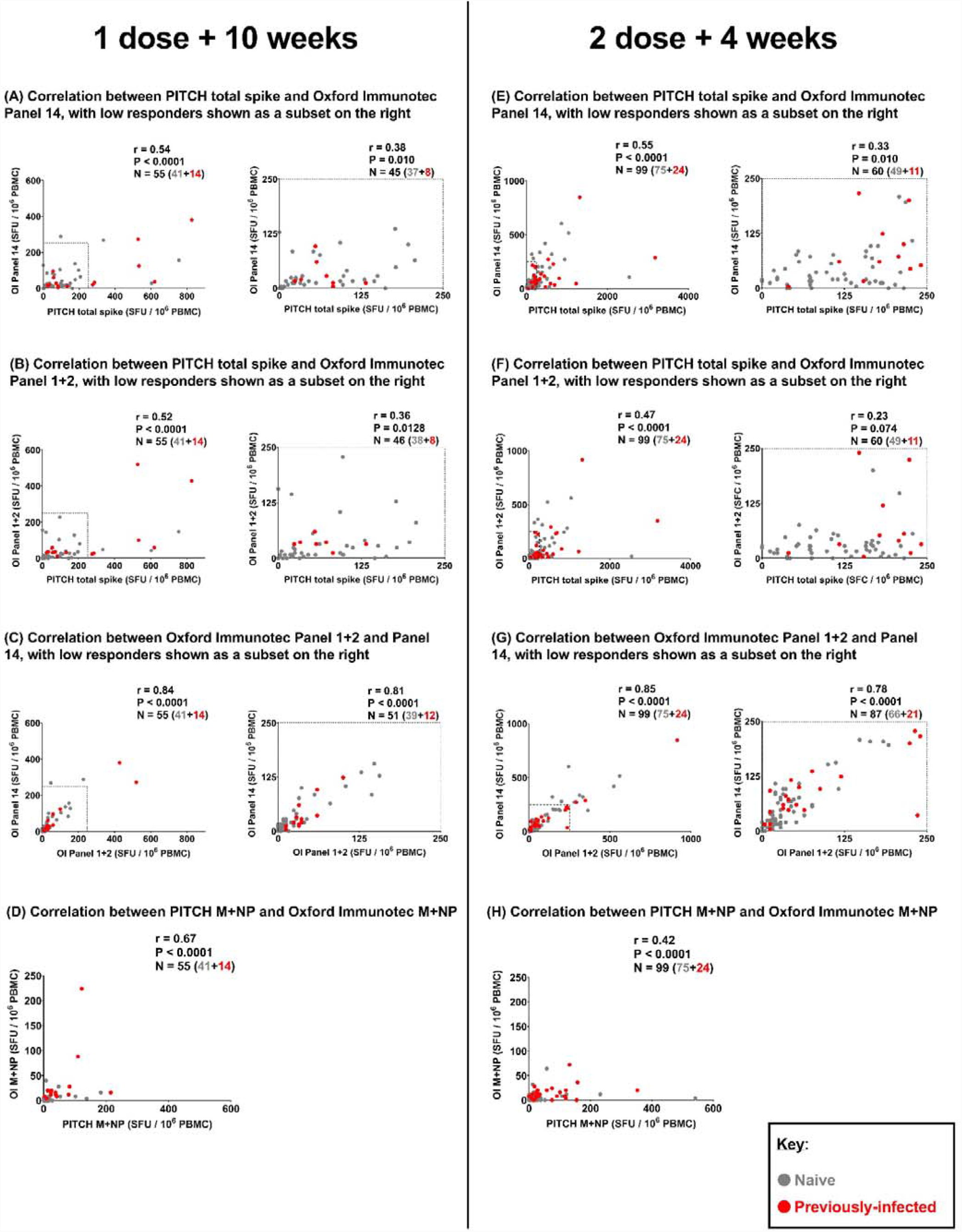
Correlation between T cell responses to SARS-CoV-2 spike and structural proteins measured by in-house PITCH ELISpot and OI T-SPOT assay. (A-C) Correlation between T cell responses to SARS-CoV-2 spike protein measured at 1 dose + 10 weeks timepoint by 3 different panels: PITCH total spike, Oxford Immunotec Panel 1+2 and Oxford Immunotec Panel 14. n=55 (n=41 naïve, n=14 previously infected). (A) Correlation between PITCH total spike and Oxford Immunotec Panel 14 at 1 dose + 10 weeks, with low responders (≤250 SFC/10^6^ PBMC) delineated by a dotted-line in the graph on the left and represented in the graph on the right (n=45 total; n=37 naïve and n=8 previously-infected). (B) Correlation between PITCH total spike and Oxford Immunotec Panel 1+2 at 1 dose + 10 weeks, with low responders (≤250 SFC/10^6^ PBMC) delineated by a dotted-line in the graph on the left and represented in the graph on the right (n=46 total; n=38 naïve and n=8 previously-infected). (C) Correlation between Oxford Immunotec Panel 1+2 and Panel 14 at 1 dose + 10 weeks, with low responders (≤250 SFC/10^6^ PBMC) delineated by a dotted-line in the graph on the left and represented in the graph on the right (n=51 total; n=39 naïve, n=12 previously-infected). (D) Correlation between T cell responses to SARS-CoV-2 membrane and nucleocapsid (M+NP) protein at 1 dose + 10 weeks timepoint measured by PITCH ELISpot and Oxford Immunotec T-SPOT. n=58 (n=41 naïve, n=14 previously infected). (E-G) Correlation between T cell responses to SARS-CoV-2 spike protein measured at 2 dose + 4 weeks timepoint by 3 different panels: PITCH total spike, Oxford Immunotec Panel 1+2 and Oxford Immunotec Panel 14. n=99 (n=75 naïve, n=24 previously infected). (E) Correlation between PITCH total spike and Oxford Immunotec Panel 14 at 2 dose + 4 weeks, with low responders (≤250 SFC/10^6^ PBMC) delineated by a dotted-line in the graph on the left and represented in the graph on the right (n=60 total; n=49 naïve and n=11 previously-infected). (F) Correlation between PITCH total spike and Oxford Immunotec Panel 1+2 at 2 dose + 4 weeks, with low responders (≤250 SFC/10^6^ PBMC) delineated by a dotted-line in the graph on the left and represented in the graph on the right (n=60 total; n=49 naïve and n=11 previously-infected). (G) Correlation between Oxford Immunotec Panel 1+2 and Panel 14 at 2 dose + 4 weeks, with low responders (≤250 SFC/10^6^ PBMC) delineated by a dotted-line in the graph on the left and represented in the graph on the right (n=87 total; n=66 naïve, n=21 previously-infected). (H) Correlation between T cell responses to SARS-CoV-2 membrane and nucleocapsid (M+NP) protein at 2 dose + 4 weeks timepoint measured by PITCH ELISpot and Oxford Immunotec T-SPOT. n=99 (n=75 naïve, n=24 previously infected). (A-H) Spearman’s r correlation was performed and two-tailed P values reported (alpha = 0.05). Infection status at time of first vaccine, as defined by available PCR and serology data: grey symbols = naïve HCWs and red symbols = HCWs previously infected with SARS-CoV-2.

Since spike-specific T cell responses measured by OI T-SPOT panels tended lower than those measured by PITCH ELISpot, we investigated whether the correlation between OI T-SPOT readouts (Panel 1+2 and Panel 14) and PITCH total spike changed when looking at low responders only. This subset was defined as having T cell responses equal to or lower than 250 SFU/10^6^ PBMCs in both panels being correlated. At both 1 dose + 10 weeks (**Figure 2A-C**) and 2 dose + 4 weeks (**Figure 2E-G**), OI spike Panels (Panel 1+2 and Panel 14) and PITCH total spike weakly correlated, ranging from r=0.23 to r=0.38. These correlations suggest that the PITCH ELISpot is more sensitive at detecting T cell responses in low responders. However, overall OI T-SPOT valuably characterises T cell responses, which is an important component of COVID-19 research as T cell and antibody responses do not always correlate (**Figure S2**).

### Defining positive responders to SARS-CoV-2 spike in both assays

We sought to investigate how positive responders could be numerically defined for each assay, and whether the proportion of positive responders differed across the three panels measuring SARS-CoV-2 spike-specific T cell responses (**Table 2**). The OI T-SPOT assay uses test wells with cell concentrations equating to 250,000 PBMCs/well, while the PITCH ELISpot assay uses 200,000 PBMCs/well. We explored positivity as defined by a cut-off of 24 SFU/10^6^ PBMCs for the OI T-SPOT and 26 SFU/10^6^ PBMCs for the PITCH SOP ELISpot. OI provided the 24 SFU/10^6^ PBMCs cut-off (based on in-house research and development defining their cut-off as 6 SFU/250,000 PBMC) whereas we calculated the 26 SFU/10^6^ PBMCs PITCH SOP ELISpot cut-off as: the mean of all negative control wells + 2 standard deviations. Using these cut-offs, we find that the percentage of positive responders measured on OI Panel 1+2 and OI Panel 14 are comparable at 1 dose + 10 weeks (55% and 49%, respectively) and 2 dose + 4 weeks (63% and 69%), whereas PITCH total spike reports a higher proportion of positive responders at both 1 dose + 10 weeks (38/55, 69%) and at 2 dose + 4 weeks (93/99, 94%). The OI assay therefore detects T cells using this cut off in a lower proportion that the PITCH ELISpot assay.

**Table 2.** Table showing the number and percentage of positive responders to SARS-CoV-2 spike at 1 dose + 10 weeks and 2 dose + 4 weeks measured across 3 different panels: Oxford Immunotec Panel 1+2 and Panel 14 and PITCH total spike. Oxford Immunotec T-SPOT assay defines the cut-off for a positive responder as 6 SFU/250,000 PBMCs, which translates to 24 SFU/10^6^ PBMCs. The PITCH ELISpot assay defines the cut-off for a positive responder as 26 SFU/10^6^ PBMCs. This cut-off is calculated from negative control wells as: (mean + 2 standard deviations). The cohort analysed here is described in Table 1 (naïve and previously infected groups are combined).

|  | n | # responders on<br>Oxford Immunotec<br>Panel 1+2 (%) | # responders on<br>Oxford Immunotec<br>Panel 14 (%) | # responders on<br>PITCH total spike (%) |
| --- | --- | --- | --- | --- |
| **Cut-off (SFU/10<sup>6</sup>**<br>**PBMCs)** | **NA** | **24** | **24** | **26** |
| **1 dose + 10**<br>**weeks** | **55** | **30**<br>**(55%)** | **27**<br>**(49%)** | **38**<br>**(69%)** |
| **2 dose + 4**<br>**weeks** | **99** | **62**<br>**(63%)** | **68**<br>**(69%)** | **93**<br>**(94%)** |

Baseline T cell response measurements with either the OI T-SPOT assay or the PITCH ELISpot assay were available in a subset of HCWs only, without matched samples (**Figure S3**). We have previously established that the PITCH ELISpot assay is highly specific with no or minimal responses in pre-pandemic samples and in individuals early in the pandemic without exposure [26]. A few individuals identified as infection naïve (no history of positive PCR test and seronegative for anti-S and anti-N antibodies) at baseline prior to vaccination (December 2020 onwards) have T cell responses to spike, and/or membrane and nucleocapsid proteins. HCWs were generally exposed to the virus in 2020, and these T cell responses may represent undiagnosed SARS-CoV-2 infection in the absence of seroconversion as previously described [26, 28].

## Discussion

The cohort included both SARS-CoV-2-naïve and previously infected healthcare workers vaccinated with either BNT162b Pfizer/BioNTech or ChAdOx1 nCoV-19 AZD1222, and receiving phlebotomy prior to 2^nd^ vaccine dose (1 dose + 10 weeks) or 4 weeks post 2^nd^ dose (2 dose + 4 weeks). T cell responses to SARS-CoV-2 spike, M and NP proteins were lower when reported by OI Panels than by PITCH ELISpot, with correlation between the assays. The OI T-SPOT assay appeared less effective at quantifying T cell responses in low responders. As OI Panel 14 and OI Panel 1+2 correlated strongly in both entire cohorts and low responders, this suggests that either readouts may be used to quantify spike-specific T cell responses. Similarly, the OI T-SPOT assay may also be used to quantify T cell responses to SARS-CoV-2 M+NP as significant differences were reported between naïve and previously-infected HCWs.

The Oxford Immunotec T-SPOT Discovery assay has been used to assess SARS-CoV-2 T cell responses in a variety of UK research settings, particularly to gain an understanding of the T cell response after vaccination. Prendecki *et al*. used the OI T-SPOT assay to monitor spike-specific T cell responses in HCWs 3 weeks after a first dose of BNT162b Pfizer/BioNTech, importantly showing that previously infected HCWs had T cell responses 10-fold higher than naïve HCWs [29]. Parry *et al*. similarly sought to characterise the spike-specific T cell responses post-vaccination, instead at 2 weeks after a second dose of BNT162b Pfizer/BioNTech and importantly in cohorts aged 80-96 years [30, 31]. While cellular responses to spike were less common than antibody responses, 63% of people had detectable T cell responses, demonstrating the utility of the OI T-SPOT assay in a cohort where T cell responses might be lower due to immunosenescence [32]. In both these studies, the OI T-SPOT assay provided rapid and crucial insight into the cellular response, accompanying characterisations of the humoral response. These findings are important as antibody and T cell responses do not always correlate [33] and the correlates of protection from SARS-CoV-2 remain to be determined. The OI T-SPOT assay provides the opportunity to study T cell responses alongside antibody responses in the absence of a T cell research laboratory, thus enabling the immune response to SARS-CoV-2 in different contexts to be determined. In addition, the OI T-SPOT assay has been utilised in large cohorts, for example the Com-Cov-2 clinical trial, characterising immune responses in participants vaccinated with different combinations of COVID-19 vaccines [34], and the UK OCTAVE study, which evaluates immune responses in vaccinated patients with immune-mediated inflammatory diseases, including cancer, inflammatory arthritis, diseases of the kidney or liver, or patients who are having a stem cell transplant [35].

## Limitations

This study is limited to healthy UK HCWs aged 21-66 years, and the T cell response after vaccination is known to be lower in some people due to ageing [31], underlying diseases and / or drug therapy leading to immunocompromise [36]. Our cohort was biased towards a higher proportion of females (63%) but larger studies have not reported sex to be of major significance for T cell responses to SARS-CoV-2 vaccines [3]. 15% of the cohort were from an ethnic minority which is reasonable representation for the UK, but further studies are needed for wider representation of all ethnicities and in other countries.

## Conclusion

Here we show that the OI T-SPOT assay is a robust method of measuring T cell responses to SARS-CoV-2 vaccination and infection, with particular benefits in national studies with relatively large cohorts. The OI T-SPOT assay offers the opportunity for evaluation at a relative scale not usually offered by T cell research laboratories in the academic sector, and in settings where a research laboratory is not available. Additional benefits include the ability of OI to receive samples up to 32 hours from blood draw (compared to 4 hours for the PITCH protocol), the rapid turnaround time of results, which are received within a week of samples being dispatched to the laboratory, and standardisation for comparing across different centres and studies. Disadvantages include the cost, the need to arrange transportation of fresh samples to the south of England, lower detection of T cell responses compared to a research laboratory, and less flexibility to customise the assay as the pandemic evolves. Further evaluation is needed of the use of the OI assay for longer follow-up periods post vaccination and in immunocompromised patients, and further development to raise its sensitivity may enhance its utility in the study of low responders. Antigen-specific interferon-gamma responses are only one of many available measures of T cell function. Overall, however the OI T-SPOT assay offers an efficient and standardised approach for researchers for comparisons across vaccine platforms, dosing approaches and research studies.

## Data Availability

The data underlying this article are available in the article and in its online supplementary material.

## Funding Statement

This work was funded by the National Core Study: Immunity (NCSi4P programme) ‘Optimal cellular assays for SARS-CoV-2 T cell, B cell and innate immunity’ and by the UK Department of Health and Social Care as part of the PITCH (Protective Immunity from T cells to Covid-19 in Health workers) Consortium, with contributions from UKRI/NIHR through the UK Coronavirus Immunology Consortium (UK-CIC), the Huo Family Foundation and The National Institute for Health Research (UKRIDHSC COVID-19 Rapid Response Rolling Call, Grant Reference Number COV19-RECPLAS).

A.A. is funded by a Wellcome Clinical Research Training Fellowship (216417/Z/19/Z). E.B. and P.K. are NIHR Senior Investigators and P.K. is funded by WT109965MA. S.J.D. is funded by an NIHR Global Research Professorship (NIHR300791). D.S. is supported by the NIHR Academic Clinical Lecturer programme in Oxford. LT is supported by a Wellcome Trust fellowship [205228/Z/16/Z]. For the purpose of Open Access, the authors have applied a CC BY public copyright licence to any Author Accepted Manuscript version arising from this submission. LT is also supported by the U.S. Food and Drug Administration Medical Countermeasures Initiative contract 75F40120C00085 and by the National Institute for Health Research Health Protection Research Unit (HPRU) in Emerging and Zoonotic Infections (NIHR200907) at University of Liverpool in partnership with Public Health England (PHE), in collaboration with Liverpool School of Tropical Medicine and the University of Oxford. LT is based at University of Liverpool.

The views expressed are those of the author(s) and not necessarily those of the NHS, the NIHR, the Department of Health and Social Care or UKHSA or the US Food and Drug Administration.

## Conflict of Interest Disclosure

S.J.D declares fees as a Scientific Advisor to the Scottish Parliament on COVID-19. D.W.E. declares lecture fees from Gilead, outside the submitted work. No other competing interests declared.

Oxford Immunotec assays were performed as part of a commercial contract and the company played no role in this report.

## Ethics Approval Statement

Participants were enrolled in the OPTIC study (GI Biobank Study 16/YH/0247, approved by the research ethics committee (REC) at Yorkshire & The Humber - Sheffield Research Ethics Committee on 29 July 2016, and amended for the OPTIC study on 8 June 2020.

## Patient Consent Statement

All participants in the study gave written, informed consent.

## Permission to Reproduce Material from Other Sources

Not applicable

## Clinical Trial Registration

Not applicable

## Acknowledgements

We are grateful to all our healthcare worker colleagues who participated in the study, and to Alex Mentzer, Lisa Fielding, Siobhan Gardiner, Anni Jämsén, and Sile Johnson for work collecting samples.

## Author Contributions

Conceptualization, S.J.D., P.K. and E.B.; Methodology, S.J.D., P.K., E.B., E.P., S.A., T.M., L.S., S.L.D., L.T., A.A., A.O. D.E., K.J., C.P.C., C.D.; Formal Analysis, E.P., S.J.D., A.O.; Data Curation, E.P, A.D., D.E.; Writing – Original Draft, E.P., S.J.D., Writing – Review & Editing, E.B., P.K., L.T.; Visualization, E.P.; Supervision, S.J.D., P.K., E.B., L.T.; Project Administration, A.D.; Funding Acquisition, P.K., S.J.D., E.B., L.T.

## PITCH Consortium

Eleanor Barnes, University of Oxford

Jeremy Chalk, University of Oxford

Susanna Dunachie, University of Oxford

Christopher Duncan, Newcastle University

Paul Klenerman, University of Oxford

Philippa Matthews, University of Oxford

Rebecca Payne, Newcastle University

Alex Richter, University of Birmingham

Thushan de Silva, University of Sheffield

Sarah Rowland-Jones, University of Sheffield

Lance Turtle, University of Liverpool

Dan Wootton, University of Liverpool

## Supplementary Figures

**Supplementary Table 1.** Table showing the median and interquartile range for T cell responses to SARS-CoV-2 spike and structural proteins measured by PITCH ELISpot and Oxford Immunotec T-SPOT assays, subdivided by timepoint after vaccination and infection status. Healthcare workers received phlebotomy at 10 weeks post 1^st^ dose (1 dose + 10 weeks) and/or at 4 weeks post 2^nd^ dose (2 dose + 4 weeks). Infection status is divided into two groups: naïve and previously infected. All reported values are measured in spot forming units/10^6^ peripheral blood mononuclear cells (SFU/ 10^6^ PBMCs). IQR denotes interquartile range.

|  | 1 dose + 10 weeks |  | 2 dose + 4 weeks |  |
| --- | --- | --- | --- | --- |
|  | Naïve<br>(n=41) | Previously infected<br>(n=14) | Naïve<br>(n=75) | Previously infected<br>(n=24) |
| Median (IQR)<br>(SFU/10<sup>6</sup> PBMCs) |  |  |  |  |
| PITCH total spike | 52 (20-138) | 107 (57-529) | 167 (75-284) | 312 (189-645) |
| Oxford Immunotec<br>Panel 1+2 | 12 (6-42) | 34 (27-70) | 28 (16-64) | 54 (32-230) |
| Oxford Immunotec<br>Panel 14 | 20 (12-70) | 30 (15-103) | 40 (16-96) | 86 (48-184) |
| PITCH M+NP | 3 (0-15) | 39 (21-90) | 7 (0-18) | 47 (22-119) |
| Oxford Immunotec<br>M+NP | 0 (0-4) | 16 (11-22) | 0 (0-4) | 12 (5-20) |

**Supplementary Figure 1.**
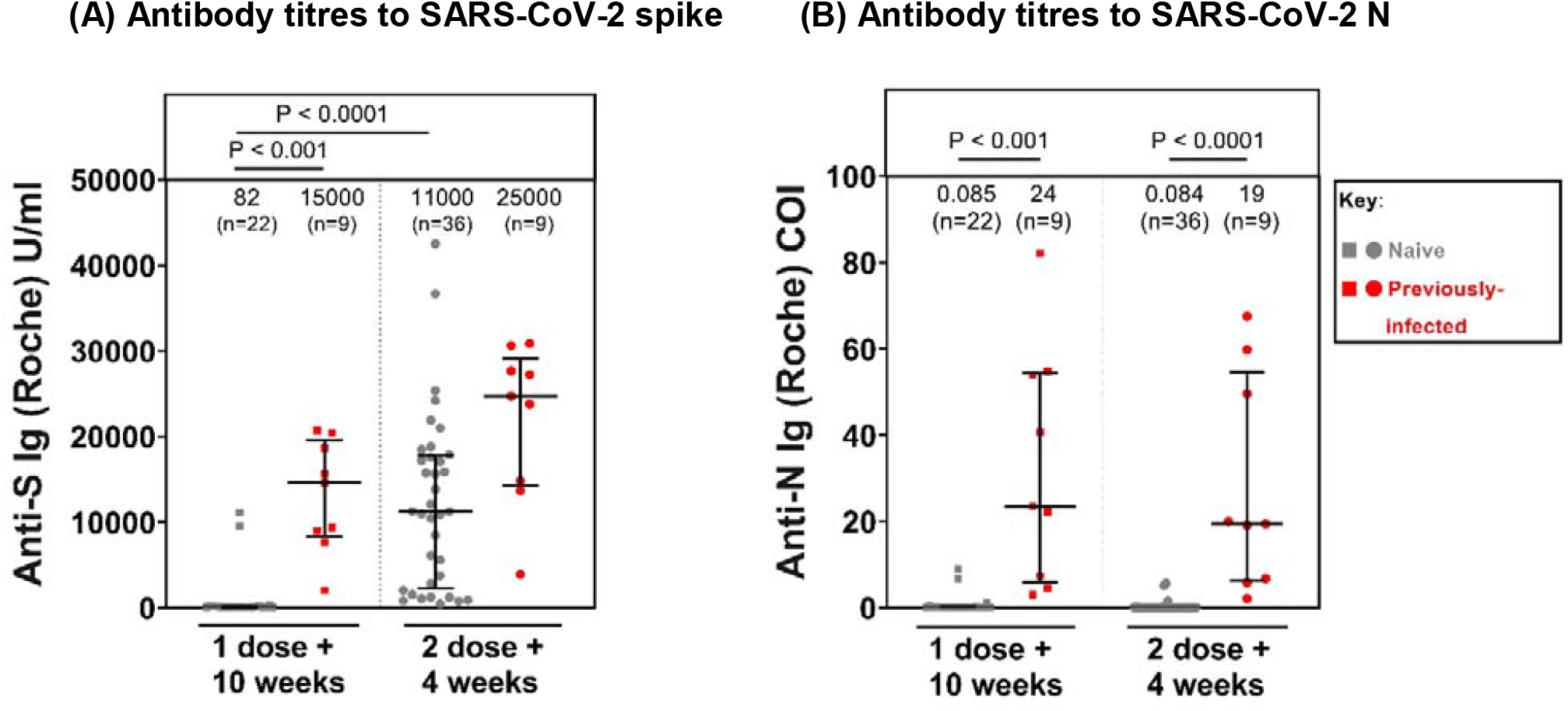
Antibody titres to SARS-CoV-2 spike and nucleocapsid protein in both naïve and previously-infected healthcare workers at 1 dose + 10 weeks and 2 dose + 4 weeks. (A) Antibody titres to SARS-CoV-2 spike protein in naïve and previously infected healthcare workers at 1 dose + 10 weeks and 2 dose + 4 weeks. (B) Antibody titres to SARS-CoV-2 nucleocapsid protein in naïve and previously infected healthcare workers at 1 dose + 10 weeks and 2 dose + 4 weeks. (A-B) Healthcare workers received phlebotomy 10 weeks post 1^st^ dose (1 dose + 10 weeks) and/or 4 weeks post 2^nd^ dose (2 dose + 4 weeks). Patients are matched for data measured by PITCH ELISpot and Oxford Immunotec T-SPOT assays. Roche assay was used to determine Antibody (Ig) titres in U/ml for anti-S and cut-off index (COI) for anti-N. At 1 dose + 10 weeks, n=22 for naïve samples and n=9 for previously-infected samples. At 2 dose + 4 weeks, n=36 for naïve samples and n=9 for previously infected samples. Kruskal-Wallis test was used to compare sample groups and determine statistical significance. Infection status at time of first vaccine, as defined by available PCR and serology data: grey symbols = naïve HCWs and red symbols = HCWs previously infected with SARS-CoV-2. Median T cell responses are stated immediately above each column and marked by a horizontal line on each column, and interquartile range is represented by error bars. The sample sizes are also indicated above each column in brackets.

**Supplementary Figure 2.**
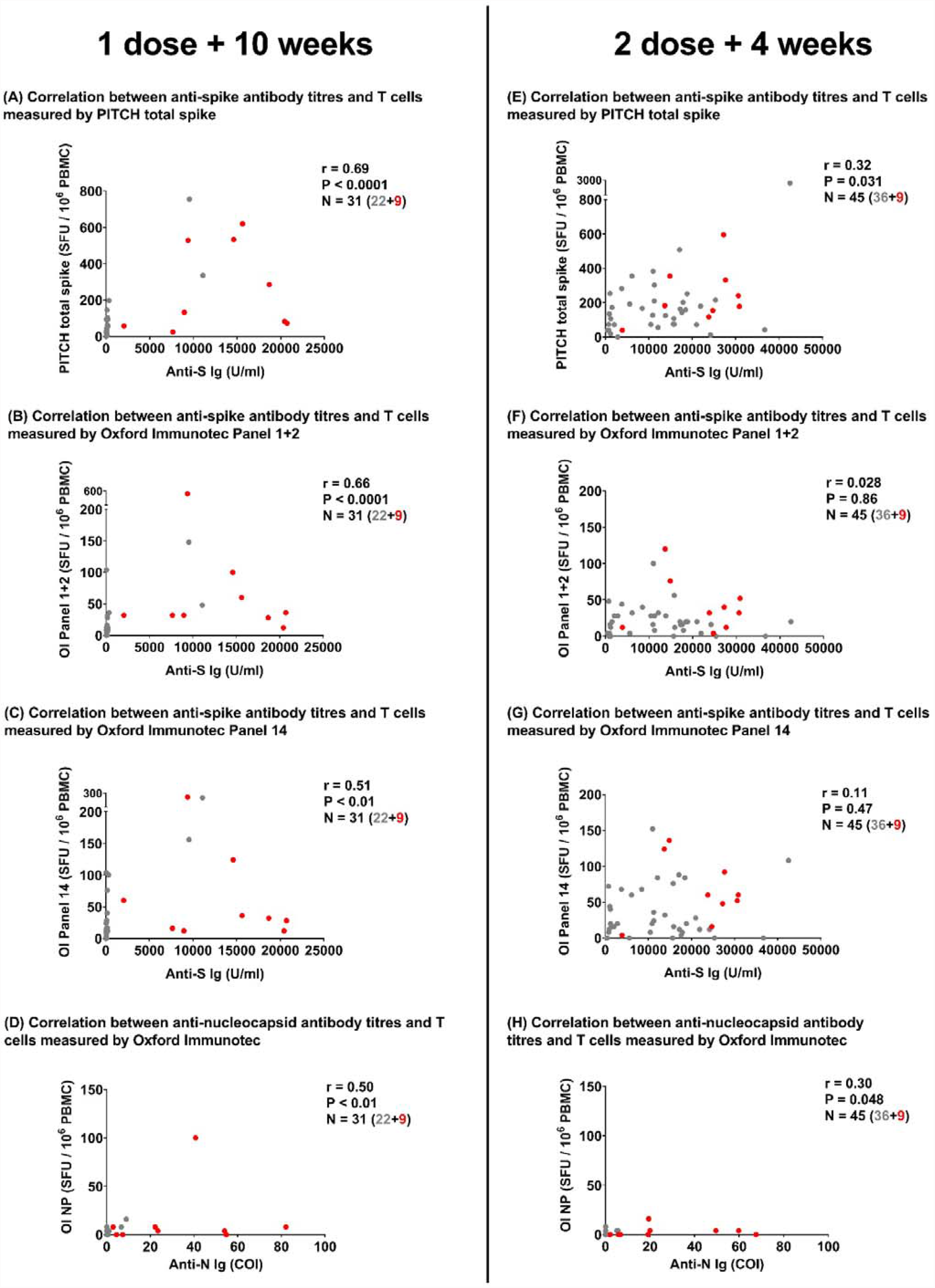
Correlation between T cell responses and Antibody titres to SARS-CoV-2 spike and nucleocapsid protein. T cell responses to spike are reported by 3 panels: PITCH total spike, Oxford Immunotec Panel 1+2 and Oxford Immunotec Panel 14. T cell responses to nucleocapsid protein are reported by Oxford Immunotec only. Roche assay was used to determine Antibody (Ig) titres in U/ml for anti-S and cut-off index (COI) for anti-N. (A-C) Correlation between antibody titres and T cell responses to SARS-CoV-2 spike protein measured at 1 dose + 10 weeks. n=31 (n=22 naïve, n=9 previously infected). (A) Correlation between SARS-CoV-2 spike antibody titres and T cell responses measured by PITCH total spike at 1 dose + 10 weeks. (B) Correlation between SARS-CoV-2 spike antibody titres and T cell responses measured by Oxford Immunotec Panel 1+2 at 1 dose + 10 weeks. (C) Correlation between SARS-CoV-2 spike antibody titres and T cell responses measured by Oxford Immunotec Panel 14 at 1 dose + 10 weeks.(D) Correlation between SARS-CoV-2 nucleocapsid antibody titres and T cell responses measured by Oxford Immunotec at 1 dose + 10 weeks. n=31 (n=22 naïve, n=9 previously infected) (E-G) Correlation between antibody titres and T cell responses to SARS-CoV-2 spike protein measured at 2 dose + 4 weeks. n=45 (n=36 naïve, n=9 previously infected). (E) Correlation between SARS-CoV-2 spike antibody titres and T cell responses measured by PITCH total spike at 2 dose + 4 weeks. (F) Correlation between SARS-CoV-2 spike antibody titres and T cell responses measured by Oxford Immunotec Panel 1+2 at 2 dose + 4 weeks. (G) Correlation between SARS-CoV-2 spike antibody titres and T cell responses measured by Oxford Immunotec Panel 14 at 2 dose + 4 weeks. (H) Correlation between SARS-CoV-2 nucleocapsid antibody titres and T cell responses measured by Oxford Immunotec at 2 dose + 4 weeks. n=45 (n=36 naïve, n=9 previously infected). Spearman’s r correlation was performed and two-tailed p values reported (alpha = 0.05). Infection status at time of first vaccine, as defined by available PCR and serology data: grey symbols = naïve HCWs and red symbols = HCWs previously infected with SARS-CoV-2.

**Supplementary Figure 3.**
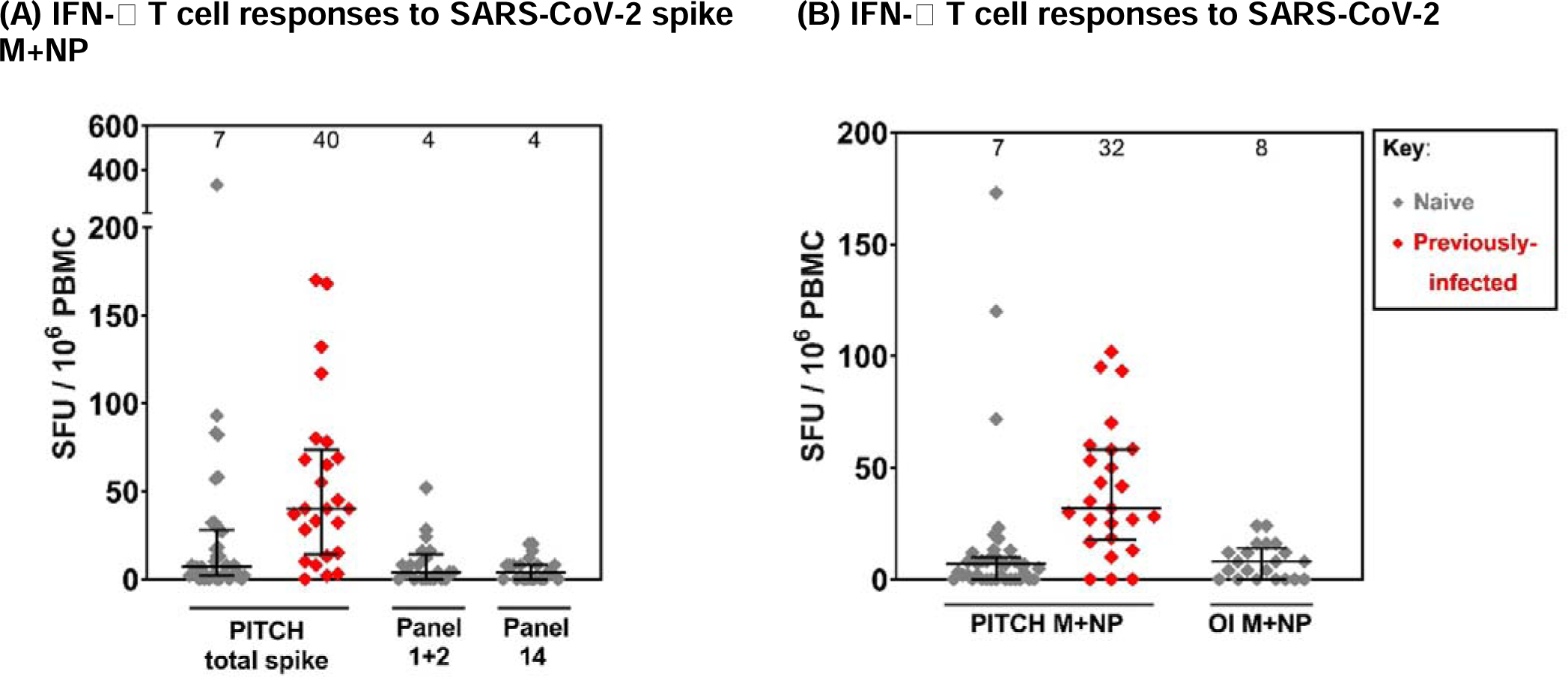
T cell responses to SARS-CoV-2 spike and structural proteins measured at baseline (pre-vaccination) by in-house PITCH ELISpot and Oxford Immunotec T-SPOT assay. Healthcare workers received phlebotomy prior to 1^st^ dose vaccination (baseline). Patients here are not matched across PITCH panels and Oxford Immunotec panels. For PITCH total spike and M+NP participants, data is matched. n=39 naïve and n=25 previously infected with SARS-CoV-2. For Oxford Immunotec Panels 1+2, 14 and M+NP, data is matched. n=21 naïve (no previously infected participants available). Grey symbols indicate naïve patients and red symbols indicate patients previously infected with SARS-CoV-2, as defined by available PCR and serology. Median T cell responses are stated immediately above each column and marked by a horizontal line on each column, and interquartile range is represented by error bars. This dataset is separate from that included in Table 1. Median age is 32 years (range: 22-72 years) with 35% and 65% of the cohort being male and female, respectively.

STROBE Statement—Checklist of items that should be included in reports of ***cohort studies***

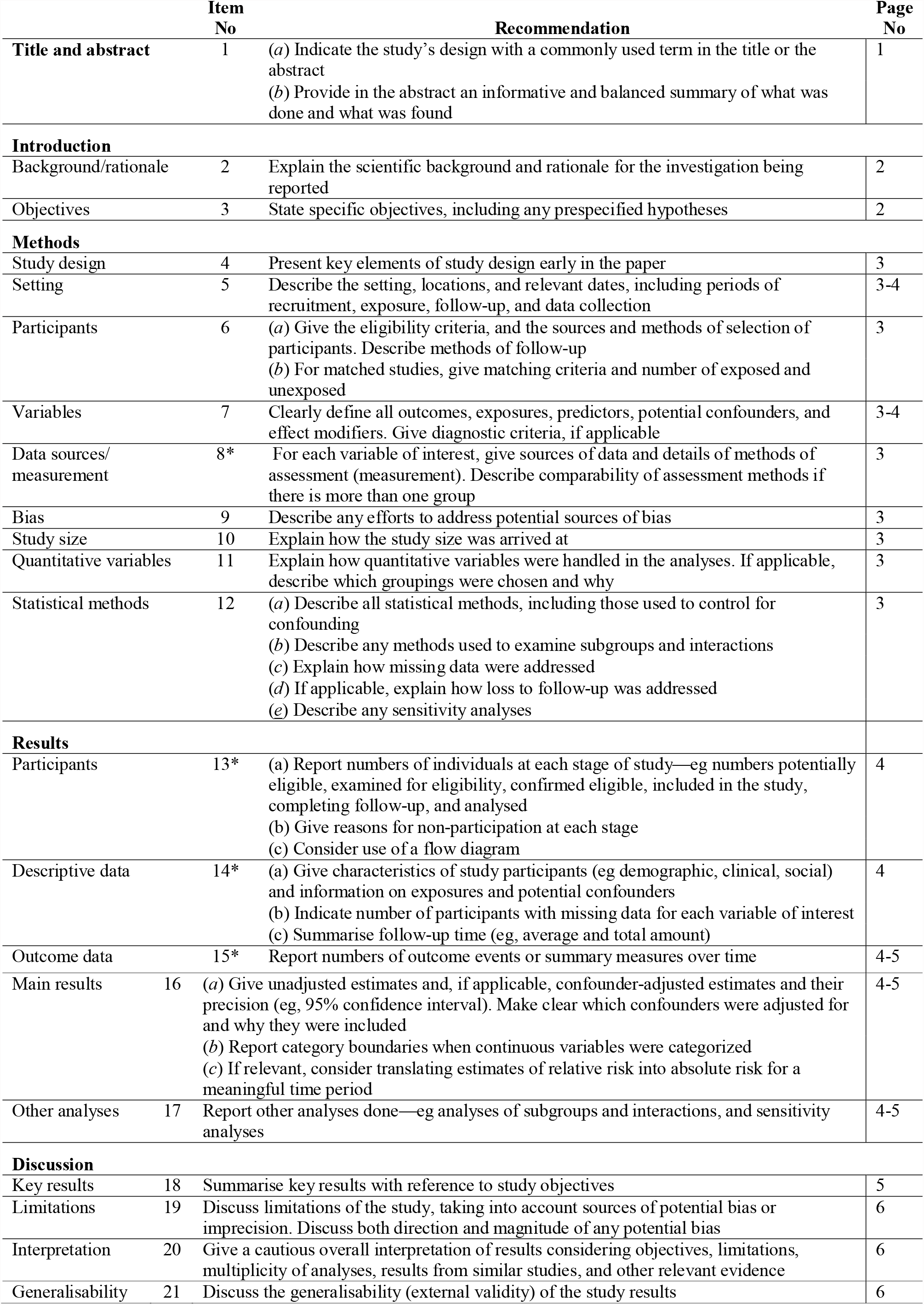
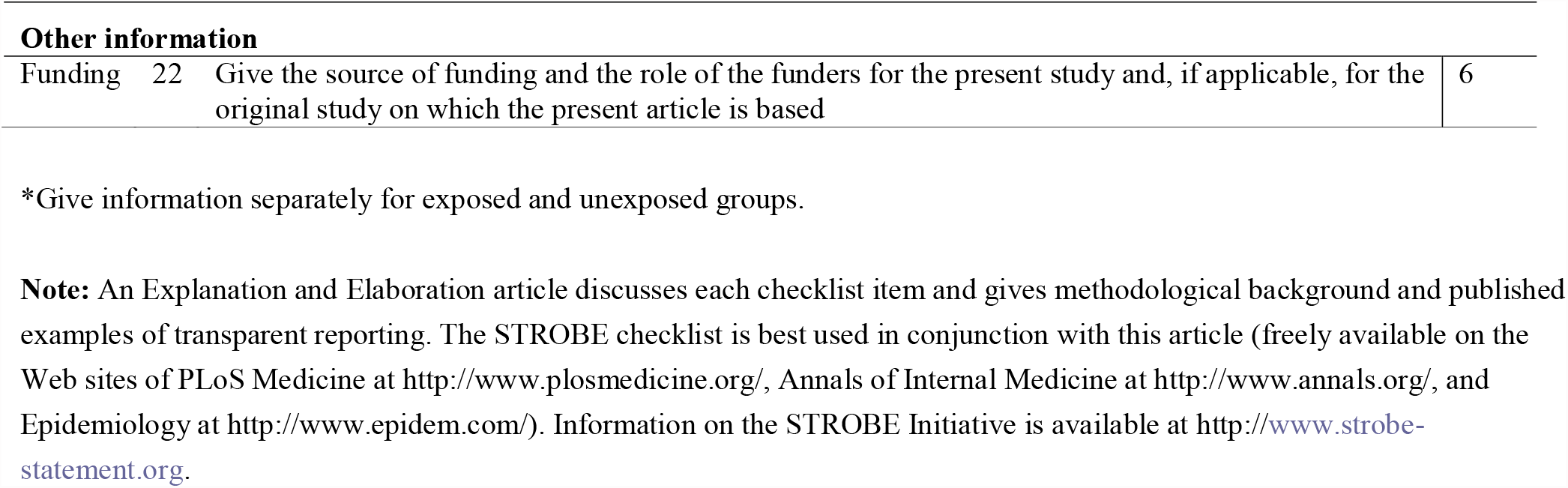

|  | Item No | Recommendation | Page No |
| --- | --- | --- | --- |
| **Title and abstract** | 1 | (a) Indicate the study's design with a commonly used term in the title or the abstract<br>(b) Provide in the abstract an informative and balanced summary of what was done and what was found | 1 |
| **Introduction** |  |  |  |
| Background/rationale | 2 | Explain the scientific background and rationale for the investigation being reported | 2 |
| Objectives | 3 | State specific objectives, including any prespecified hypotheses | 2 |
| **Methods** |  |  |  |
| Study design | 4 | Present key elements of study design early in the paper | 3 |
| Setting | 5 | Describe the setting, locations, and relevant dates, including periods of recruitment, exposure, follow-up, and data collection | 3-4 |
| Participants | 6 | (a) Give the eligibility criteria, and the sources and methods of selection of participants. Describe methods of follow-up<br>(b) For matched studies, give matching criteria and number of exposed and unexposed | 3 |
| Variables | 7 | Clearly define all outcomes, exposures, predictors, potential confounders, and effect modifiers. Give diagnostic criteria, if applicable | 3-4 |
| Data sources/measurement | 8* | For each variable of interest, give sources of data and details of methods of assessment (measurement). Describe comparability of assessment methods if there is more than one group | 3 |
| Bias | 9 | Describe any efforts to address potential sources of bias | 3 |
| Study size | 10 | Explain how the study size was arrived at | 3 |
| Quantitative variables | 11 | Explain how quantitative variables were handled in the analyses. If applicable, describe which groupings were chosen and why | 3 |
| Statistical methods | 12 | (a) Describe all statistical methods, including those used to control for confounding<br>(b) Describe any methods used to examine subgroups and interactions<br>(c) Explain how missing data were addressed<br>(d) If applicable, explain how loss to follow-up was addressed<br>(e) Describe any sensitivity analyses | 3 |
| **Results** |  |  |  |
| Participants | 13* | (a) Report numbers of individuals at each stage of study—eg numbers potentially eligible, examined for eligibility, confirmed eligible, included in the study, completing follow-up, and analysed<br>(b) Give reasons for non-participation at each stage<br>(c) Consider use of a flow diagram | 4 |
| Descriptive data | 14* | (a) Give characteristics of study participants (eg demographic, clinical, social) and information on exposures and potential confounders<br>(b) Indicate number of participants with missing data for each variable of interest<br>(c) Summarise follow-up time (eg, average and total amount) | 4 |
| Outcome data | 15* | Report numbers of outcome events or summary measures over time | 4-5 |
| Main results | 16 | (a) Give unadjusted estimates and, if applicable, confounder-adjusted estimates and their precision (eg, 95% confidence interval). Make clear which confounders were adjusted for and why they were included<br>(b) Report category boundaries when continuous variables were categorized<br>(c) If relevant, consider translating estimates of relative risk into absolute risk for a meaningful time period | 4-5 |
| Other analyses | 17 | Report other analyses done—eg analyses of subgroups and interactions, and sensitivity analyses | 4-5 |
| **Discussion** |  |  |  |
| Key results | 18 | Summarise key results with reference to study objectives | 5 |
| Limitations | 19 | Discuss limitations of the study, taking into account sources of potential bias or imprecision. Discuss both direction and magnitude of any potential bias | 6 |
| Interpretation | 20 | Give a cautious overall interpretation of results considering objectives, limitations, multiplicity of analyses, results from similar studies, and other relevant evidence | 6 |
| Generalisability | 21 | Discuss the generalisability (external validity) of the study results | 6 |

| Other information |  |  |  |
| --- | --- | --- | --- |
| Funding | 22 | Give the source of funding and the role of the funders for the present study and, if applicable, for the original study on which the present article is based | 6 |

\*Give information separately for exposed and unexposed groups.
**Note:** An Explanation and Elaboration article discusses each checklist item and gives methodological background and published examples of transparent reporting. The STROBE checklist is best used in conjunction with this article (freely available on the Web sites of PLoS Medicine at http://www.plosmedicine.org/, Annals of Internal Medicine at http://www.annals.org/, and Epidemiology at http://www.epidem.com/). Information on the STROBE Initiative is available at http://www.strobe-statement.org.

## References

1. Lumley SF, O’Donnell D, Stoesser NE, Matthews PC, Howarth A, Hatch SB, Eyre DW. Antibody Status and Incidence of SARS-CoV-2 Infection in Health Care Workers. New England Journal of Medicine 2020; 384:533–40. 10.1056/NEJMoa2034545.

2. Hall VJ, Foulkes S, Charlett A, Atti A, Monk EJM, Simmons R, Hopkins S. SARS-CoV-2 infection rates of antibody-positive compared with antibody-negative health-care workers in England: a large, multicentre, prospective cohort study (SIREN). Lancet 2021; 397:1459–69 10.1016/s0140-6736(21)00675-9.

3. Payne RP, Longet S, Austin JA, Skelly DT, Dejnirattisai W, Adele S, Consortium P. Immunogenicity of standard and extended dosing intervals of BNT162b2 mRNA vaccine. Cell 2021; 184:5699–714.e11. 10.1016/j.cell.2021.10.011.

4. Earle KA, Ambrosino DM, Fiore-Gartland A, Goldblatt D, Gilbert PB, Siber GR, Plotkin SA. Evidence for antibody as a protective correlate for COVID-19 vaccines. Vaccine 2021; 39:4423–8. https://doi.org/10.1016/j.vaccine.2021.05.063.

5. Feng S, Phillips DJ, White T, Sayal H, Aley PK, Bibi S, the Oxford CVTG. Correlates of protection against symptomatic and asymptomatic SARS-CoV-2 infection. medRxiv 2021:2021.06.21.21258528. 10.1101/2021.06.21.21258528.

6. Khoury DS, Cromer D, Reynaldi A, Schlub TE, Wheatley AK, Juno JA, Davenport MP. What level of neutralising antibody protects from COVID-19? medRxiv 2021:DOI: 2021.03.09.21252641. 10.1101/2021.03.09.21252641.

7. Addetia A, Crawford Katharine HD, Dingens A, Zhu H, Roychoudhury P, Huang M-L, McAdam Alexander J. Neutralizing Antibodies Correlate with Protection from SARS-CoV-2 in Humans during a Fishery Vessel Outbreak with a High Attack Rate. Journal of Clinical Microbiology; 58:e02107–20. 10.1128/JCM.02107-20.

8. Moore PL, Moyo-Gwete T, Hermanus T, Kgagudi P, Ayres F, Makhado Z, Gray G. Neutralizing antibodies elicited by the Ad26.COV2.S COVID-19 vaccine show reduced activity against 501Y.V2 (B.1.351), despite protection against severe disease by this variant. bioRxiv 2021:DOI: 2021.06.09.447722. 10.1101/2021.06.09.447722.

9. McMahan K, Yu J, Mercado NB, Loos C, Tostanoski LH, Chandrashekar A, Barouch DH. Correlates of protection against SARS-CoV-2 in rhesus macaques. Nature 2021; 590:630–4. 10.1038/s41586-020-03041-6.

10. Rydyznski Moderbacher C, Ramirez SI, Dan JM, Grifoni A, Hastie KM, Weiskopf D, Crotty S. Antigen-Specific Adaptive Immunity to SARS-CoV-2 in Acute COVID-19 and Associations with Age and Disease Severity. Cell 2020; 183:996–1012.e19. 10.1016/j.cell.2020.09.038.

11. Payne RP, Longet S, Austin JA, Skelly DT, Dejnirattisai W, Adele S, Zawia AAT. Immunogenicity of standard and extended dosing intervals of BNT162b2 mRNA vaccine. Cell 2021; 184:5699–714.e11. https://doi.org/10.1016/j.cell.2021.10.011.

12. Skelly DT, Harding AC, Gilbert-Jaramillo J, Knight ML, Longet S, Brown A, Group CMP-C. Two doses of SARS-CoV-2 vaccination induce robust immune responses to emerging SARS-CoV-2 variants of concern. Nature Communications 2021; 12:5061 10.1038/s41467-021-25167-5.

13. Keeton R, Tincho MB, Ngomti A, Baguma R, Benede N, Suzuki A, Riou C. SARS-CoV-2 spike T cell responses induced upon vaccination or infection remain robust against Omicron. medRxiv 2021:2021.12.26.21268380. 10.1101/2021.12.26.21268380.

14. Tarke A, Coelho CH, Zhang Z, Dan JM, Yu ED, Methot N, Sette A. SARS-CoV-2 vaccination induces immunological memory able to cross-recognize variants from Alpha to Omicron. bioRxiv 2021:2021.12.28.474333. 10.1101/2021.12.28.474333.

15. Yu G, Curtis C, Alba G, Thomas M, Julia N, Anna O, … Marcus B. Research Square 2022. 10.21203/rs.3.rs-1217466/v1.

16. GeurtsvanKessel CH, Geers D, Schmitz KS, Mykytyn AZ, Lamers MM, Bogers S, de Vries RD. Divergent SARS CoV-2 Omicron-specific T-and B-cell responses in COVID-19 vaccine recipients. medRxiv 2021:2021.12.27.21268416. 10.1101/2021.12.27.21268416.

17. Madelon N, Heikkilä N, Sabater Royo I, Fontannaz P, Breville G, Lauper K, Eberhardt CS. Omicron-specific cytotoxic T-cell responses are boosted following a third dose of mRNA COVID-19 vaccine in anti-CD20-treated multiple sclerosis patients. medRxiv 2021:2021.12.20.21268128. 10.1101/2021.12.20.21268128.

18. De Marco L, D’Orso S, Pirronello M, Verdiani A, Termine A, Fabrizio C, Santa Lucia Foundation RI. Preserved T cell reactivity to the SARS-CoV-2 Omicron variant indicates continued protection in vaccinated individuals. bioRxiv 2021:2021.12.30.474453. 10.1101/2021.12.30.474453.

19. Liu J, Chandrashekar A, Sellers D, Barrett J, Lifton M, McMahan K, Barouch DH. Vaccines Elicit Highly Cross-Reactive Cellular Immunity to the SARS-CoV-2 Omicron Variant. medRxiv 2022:2022.01.02.22268634. 10.1101/2022.01.02.22268634.

20. Karlsson AC, Martin JN, Younger SR, Bredt BM, Epling L, Ronquillo R, Sinclair E. Comparison of the ELISPOT and cytokine flow cytometry assays for the enumeration of antigen-specific T cells. J Immunol Methods 2003; 283:141–53.

21. Tanguay S, Killion JJ. Direct comparison of ELISPOT and ELISA-based assays for detection of individual cytokine-secreting cells. Lymphokine Cytokine Res 1994; 13:259–63.

22. Angyal A, Longet S, Moore SC, Payne RP, Harding A, Tipton T, de Silva TI. T-cell and antibody responses to first BNT162b2 vaccine dose in previously infected and SARS-CoV-2-naive UK health-care workers: a multicentre prospective cohort study. Lancet Microbe 2021 10.1016/s2666-5247(21)00275-5.

23. Nussenzweig V, Nussenzweig RS. Circumsporozoite proteins of malaria parasites. Cell 1985; 42:401–3.

24. Whitworth HS, Scott M, Connell DW, Dongés B, Lalvani A. IGRAs – The gateway to T cell based TB diagnosis. Methods 2013; 61:52–62. https://doi.org/10.1016/j.ymeth.2012.12.012.

25. McConkey SJ, Reece WH, Moorthy VS, Webster D, Dunachie S, Butcher G, Hill AV. Enhanced T-cell immunogenicity of plasmid DNA vaccines boosted by recombinant modified vaccinia virus Ankara in humans. Nat Med 2003; 9:729–35. 10.1038/nm881.

26. Ogbe A, Kronsteiner B, Skelly DT, Pace M, Brown A, Adland E, Oxford Protective T Cell Immunology for Covid-Clinical Team. T cell assays differentiate clinical and subclinical SARS-CoV-2 infections from cross-reactive antiviral responses. Nat Commun 2021; 12:2055. https://www.nature.com/articles/s41467-021-21856-3.

27. T cell testing in SARS-CoV-2. http://www.tspotdiscovery.com/wp-content/uploads/sites/4/2020/11/P432-UK-WP-MPN475-0001-V2-T-SPOT-Discovery-White-Paper-DIGITAL-READY-06Nov20.pdf, date accessed: 26th January 2022.

28. Swadling L, Diniz MO, Schmidt NM, Amin OE, Chandran A, Shaw E, Investigators CO. Pre-existing polymerase-specific T cells expand in abortive seronegative SARS-CoV-2. Nature 2022; 601:110–7. 10.1038/s41586-021-04186-8.

29. Prendecki M, Clarke C, Brown J, Cox A, Gleeson S, Guckian M, Willicombe M. Effect of previous SARS-CoV-2 infection on humoral and T-cell responses to single-dose BNT162b2 vaccine. The Lancet 2021; 397:1178–81. https://doi.org/10.1016/S0140-6736(21)00502-X.

30. Parry H, Tut G, Bruton R, Faustini S, Stephens C, Saunders P, Moss P. mRNA vaccination in people over 80 years of age induces strong humoral immune responses against SARS-CoV-2 with cross neutralization of P.1 Brazilian variant. Elife 2021; 10. 10.7554/eLife.69375.

31. Parry H, Bruton R, Stephens C, Brown K, Amirthalingam G, Otter A, Moss P. Differential immunogenicity of BNT162b2 or ChAdOx1 vaccines after extended-interval homologous dual vaccination in older people. Immun Ageing 2021; 18:34. 10.1186/s12979-021-00246-9.

32. Bektas A, Schurman SH, Sen R, Ferrucci L. Human T cell immunosenescence and inflammation in aging. J Leukoc Biol 2017; 102:977–88. 10.1189/jlb.3RI0716-335R.

33. Payne RP, Longet S, Austin JA, Skelly DT, Dejnirattisai W, Adele S, Dunachie S. Immunogenicity of standard and extended dosing intervals of BNT162b2 mRNA vaccine. Cell 2021; 184:5699–714.e11. 10.1016/j.cell.2021.10.011.

34. Liu X, Shaw RH, Stuart ASV, Greenland M, Aley PK, Andrews NJ, Snape MD. Safety and immunogenicity of heterologous versus homologous prime-boost schedules with an adenoviral vectored and mRNA COVID-19 vaccine (Com-COV): a single-blind, randomised, non-inferiority trial. Lancet 2021; 398:856–69. 10.1016/s0140-6736(21)01694-9.

35. Kearns P, Siebert S, Willicombe M, Gaskell C, Kirkham A, Pirrie S, McInnes I. Examining the Immunological Effects of COVID-19 Vaccination in Patients with Conditions Potentially Leading to Diminished Immune Response Capacity – The OCTAVE Trial. Lancet preprint 2021. http://dx.doi.org/10.2139/ssrn.3910058.

36. Prendecki M, Thomson T, Clarke CL, Martin P, Gleeson S, De Aguiar RC, Imperial Renal C-vsgicwtOSC. Immunological responses to SARS-CoV-2 vaccines in kidney transplant recipients. Lancet (London, England) 2021; 398:1482–4. 10.1016/S0140-6736(21)02096-1.

